# The validation and clinical applicability of angiography-derived assessment of coronary microcirculatory resistance: a [^15^O]H_2_O PET study

**DOI:** 10.1101/2023.12.05.23299545

**Authors:** Ruurt A. Jukema, Pieter G Raijmakers, Masahiro Hoshino, Roel S. Driessen, Pepijn A. van Diemen, Juhani Knuuti, Teemu Maaniitty, Jos Twisk, Rolf A. Kooistra, Janny Timmer, Johan H.C. Reiber, Pim van der Harst, Maarten J. Cramer, Tim van der Hoef, Paul Knaapen, Ibrahim Danad

## Abstract

**Background:** The introduction of wire-free microcirculatory resistance index from functional angiography (angio-IMR) promises swift detection of coronary microvascular dysfunction, however it has not been properly validated. We sought to validate angio-IMR against invasive IMR and PET derived microvascular resistance (MVR). Moreover, we studied if angio-IMR could aid in the detection of ischemia with non-obstructive coronary arteries (INOCA).

**Methods:** In this investigator-initiated study symptomatic patients underwent [^15^O]H_2_O positron emission tomography (PET) and invasive angiography with 3-vessel fractional flow reserve (FFR). Invasive IMR was measured in 40 patients. Angio-IMR and QFR were computed retrospectively. MVR was defined as the ratio of mean distal coronary pressure to PET derived coronary flow. PET and QFR/angio-IMR analyses were performed by blinded core labs. The right coronary artery was excluded.

**Results:** A total of 211 patients (mean age 61±9, 148 (70%) male) with 312 vessels with successful angio-IMR analyses were included. Angio-IMR correlated moderately with invasive IMR (r=0.48, p<0.01), whereas no correlation was found between angio-IMR and MVR (r=-0.07, p=0.25). Angio-IMR did not differ for vessels without obstructive coronary artery disease (CAD) (FFR-) but with reduced stress perfusion (PET+) compared to vessels without obstructive CAD (FFR-) with normal stress perfusion (PET-) (median 28.19 IQR 20.42 – 38.99 vs 31.67 IQR 23.47 – 40.63, p=0.40).

**Conclusion:** Angio-IMR correlated moderately with invasively measured IMR, whereas angio-IMR did not correlate with PET derived MVR. Moreover, angio-IMR was similar in patients without obstructive CAD, irrespective of ischaemia status, hampering the identification of INOCA.

## Introduction

Chronic ischemic heart disease is a multifactorial entity that is caused by either epicardial atherosclerotic coronary artery disease (CAD), coronary microvascular dysfunction (CMD) or a combination thereof.(1) Traditionally, the management of symptomatic patients is focussed on slowing the progression of atherosclerosis through medication and the detection of obstructive CAD. However, a large portion of patients referred for invasive coronary angiography (ICA) does not have obstructive CAD and the prevalence of post percutaneous coronary intervention (PCI) angina ranges from 20–40%. (2, 3) Fractional flow reserve (FFR) is used in the cathlab to assess vessel-specific ischemia, but the microvasculature remains unassessed by this epicardial measure. (4, 5) The interplay between CAD as assed by FFR and CMD has gained attention, especially since a CMD tailored medical treatment may lead to an improved quality of life. (2, 6) Yet, invasive resistance measurements are time consuming and therefore not routinely performed. Computed measurements of coronary and microvascular function hold the promise to differentiate between CAD and CMD within a single analysis. Quantitative flow ratio (QFR) has been shown to accurately predict the epicardial significance of CAD and a QFR-based treatment appears safe. (7, 8) Furthermore, the introduction of wire-free microcirculatory resistance index from functional angiography -angio-IMR-provides a direct overview of epicardial (QFR) and microvascular disease status (angio-IMR). (9, 10) Directly after introduction of angio-IMR the practical applicability was studied, but angio-IMR has yet to be validated against a golden-standard index of microvascular resistance (MVR). As such, the first aim is to validate angio-IMR against traditional invasive resistance indices and [^15^O]H_2_O PET derived microvascular resistance. The second aim is to investigate whether angio-IMR can improve the detection of ischaemia with non-obstructive coronary arteries (INOCA) in the cathlab.

## Methods

### Patient selection

This is a sub study of the Comparison of Coronary CT Angiography, SPECT, PET, and Hybrid Imaging for Diagnosis of Ischemic Heart Disease Determined by Fractional Flow Reserve (PACIFIC 1) and Functional stress imaging to predict abnormal coronary fractional flow reserve: the PACIFIC 2 study (PACIFIC 2), which were prospective clinical single-centre, head-to-head comparative studies conducted from 2012 to 2020, at the Amsterdam UMC, VU University Medical Center, Amsterdam, the Netherlands. (11, 12) All patients were suspected of having obstructive coronary artery disease (CAD) and underwent a 2-week protocol in which patients underwent [^15^O]H_2_O PET prior to invasive coronary angiography (ICA) with routine 3 vessel invasive FFR interrogation. This PACIFIC post-hoc analysis included patients with angio-IMR and invasive IMR or PET perfusion imaging. The study complied with the Declaration of Helsinki. The study protocol was approved by the VUmc Medical Ethics Review Committee, and all patients provided written informed consent.

### PET

The PET scans were performed on a hybrid PET/CT device (Philips Gemini TF 64 or Ingenuity TF 128, Philips Healthcare, Best, The Netherlands). The scanning protocol has been described in detail previously.(13) Briefly, a 6-minute dynamic scan protocol commencing simultaneously with an injection of 370 MBq [^15^O]H_2_O during resting and adenosine (140 μg/kg/min) induced hyperemic conditions. The dynamic scan sequence was followed by a low-dose CT-scan for attenuation correction. Patients were instructed to refrain from the intake of xanthine or caffeine 24 hours prior to the PET. Peripheral arterial pressure was measured 3 minutes into scanning.

### ICA and FFR

ICA was performed according to standard clinical protocols. (14) Patients were instructed to refrain from the intake of xanthine or caffeine 24 hours prior to the ICA. All major coronary arteries were routinely interrogated by FFR irrespective of stenosis severity and imaging results, except for occluded vessels or subtotal lesions with a diameter stenosis (DS) ≥90%. To induce maximal coronary hyperemia, adenosine was administered intracoronary as a 150 μg bolus or intravenously (140 μg/kg/min). FFR was calculated as the ratio of mean distal intracoronary to aortic guiding pressure during hyperemia. A FFR ≤0.80 was considered abnormal. (5) In 40 PACIFIC 1 patients invasive IMR was measured. A pressure/temperature sensor-tipped guidewire (PressureWire X wired, Abbott, Chicago, Illinois) was advanced in the distal third part of the coronary artery and connected to a RadiAnalyzer interface (Abbott). The guiding catheter was then flushed with saline. Next, 3 ml of saline at room temperature was rapidly injected through the guiding catheter. This process was repeated twice, yielding 3 thermodilution curves. Operators were encouraged to replace discordant values by repeating injections. After baseline measurements, hyperemia was induced through intravenous infusion of adenosine using the same protocol as during PET imaging. During steady-state maximum hyperemia 3 more thermodilution curves were acquired. IMR was calculated as the product of distal hyperemic pressure and mean hyperemic transit time.

### QFR and angio-IMR

First, QFR was computed. Two end-diastolic frames at least 25° apart from the coronary of interest were used to reconstruct a 3D-model of the coronary artery. The reference diameter of the vessel was constructed by marking healthy segments of the coronary artery preferably proximally and distally of a lesion of interest. Lesion length and diameter stenosis (%) were extracted from the 3D model. Contrast frame counting during resting conditions of the analysed artery was performed to obtain an estimated contrast flow velocity which is converted into a virtual hyperemic flow velocity. Subsequently contrast QFR is computed by the software package. If contrast frame counting was not possible fixed QFR was used. Secondly, the mean rest arterial pressure was calculated (the sum of all invasive arterial rest pressures divided by the amount of measurements) and angio-IMR was computed. The formula used to compute angio-IMR was described earlier by Mejia-Renteria et al. and it incorporates arterial rest pressure, QFR, vessel length and contrast flow velocity.(10) If contrast flow counting QFR was not possible and fixed QFR was used as alternative angio-IMR could not be computed. QFR analyses were post-hoc performed by a blinded core laboratory (ClinFact Medis Specials, Leiden, the Netherlands) using the QAngio XA 3D/QFR 2.1-research software package (Medis Medical Imaging Systems, Leiden, the Netherlands). Patients with chronic total occlusions were excluded for angio-IMR computation.

### Data analysis

Parametric PET images of quantitative hyperemic myocardial blood flow (hMBF) in ml/min/g were generated for each of the 17 left ventricle segments according to the standard American Heart association model and quantitatively analysed by an independent core laboratory (Turku University Hospital Finland). (15) Regional hMBF of each coronary territory was defined as mean hyperaemic MBF of the entire vascular territory in the absence of a perfusion defect or as the mean hMBF of the perfusion defect (≥2 adjacent segments with a hyperaemic MBF ≤2.3 ml/min/g) when present. (16) Regional hMBF was used to define ischaemia because of its greater ability to identify ischaemia and better prognostic value compared to coronary flow reserve (CFR) using [^15^O]H_2_O as tracer. (16, 17) MVR was defined as the ratio of mean distal coronary pressure to coronary flow. Mean distal coronary pressure was estimated by multiplying peripheral mean arterial pressure during PET with FFR of the corresponding vessels. Regional hMBF of the corresponding vessels was used to define coronary flow. The right coronary artery was excluded from analysis to minimize the influence of myocardial mass.

### Statistical analysis

Continuous variables are expressed as mean ± SD or median (interquartile range) where appropriate. Categorical variables are presented as frequencies with percentages. Normality of data was examined by means of QQ-plots and histograms. The relation between angio-IMR and invasive IMR or MVR was assessed by Spearman’s correlation coefficient. Agreement between angio-IMR and invasive IMR or MVR was assessed using Bland-Altman analyses. Analyses concerning the correlation between angio-IMR and MVR were stratified for vessels with or without obstructive CAD using 3D quantitative coronary angiography (QCA). The Kruskal-Wallis test was used to assess differences in FFR, QFR, 3D QCA and angio-IMR values across groups stratified for FFR/QFR and PET results. If the Kruskal Wallis test was significant, post-hoc pairwise comparisons between patient categories were performed using the Mann-Whitney U test with Bonferroni correction. Means of hMBF, CFR and MVR were compared using an one way analysis of variance (ANOVA). A p-value <0.05 was considered statistically significant. All analyses were performed using IBM SPSS (SPSS Statistics 26, IBM, Armonk, New York).

## Results

### Study population

The final study population consisted of 211 patients with 312 successful angio-IMR analyses. Baseline characteristics are presented in table 1. Angiographic and PET characteristics stratified for FFR and PET results are depicted in table 2. The study flow chart is shown in figure 1 and outlines the excluded patients and vessels. The computation of angio-IMR was feasible in vessels with successful contrast frame counting QFR analysis. As such, vessels without successful QFR (n=355) were excluded. Of 439 vessels with successful QFR analysis an additional 87 vessels were excluded because of the utilization of fixed flow QFR or missing arterial rest pressure. Detailed flow charts for failed QFR computation have been published before. (8, 18) A skewed distribution was observed for FFR (0.89, CI: 0.81-0.96), QFR (0.93, CI: 0.80–0.99) and angio-IMR (27.74 CI: 20.85 –38.02). Angio-IMR and MVR were available in 309 vessels, whereas angio-IMR and invasive IMR were available in 29 vessels (figure 1).

**Table 1.**
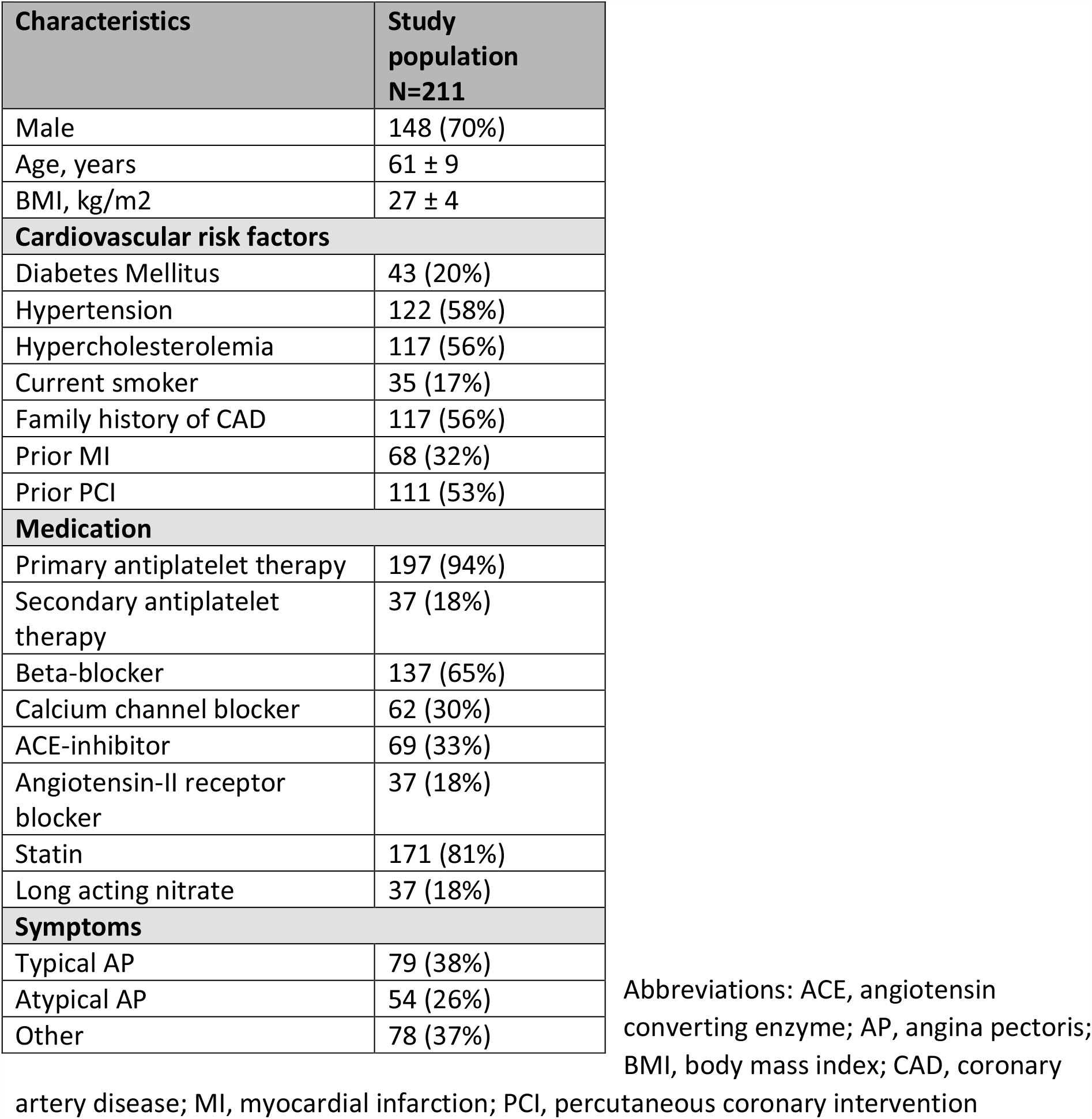
Baseline characteristics; mean ± SD or N(%)

| Characteristics | Study population<br>N=211 |
| --- | --- |
| Male | 148 (70%) |
| Age, years | 61 ± 9 |
| BMI, kg/m<sup>2</sup> | 27 ± 4 |
| **Cardiovascular risk factors** |  |
| Diabetes Mellitus | 43 (20%) |
| Hypertension | 122 (58%) |
| Hypercholesterolemia | 117 (56%) |
| Current smoker | 35 (17%) |
| Family history of CAD | 117 (56%) |
| Prior MI | 68 (32%) |
| Prior PCI | 111 (53%) |
| **Medication** |  |
| Primary antiplatelet therapy | 197 (94%) |
| Secondary antiplatelet therapy | 37 (18%) |
| Beta-blocker | 137 (65%) |
| Calcium channel blocker | 62 (30%) |
| ACE-inhibitor | 69 (33%) |
| Angiotensin-II receptor blocker | 37 (18%) |
| Statin | 171 (81%) |
| Long acting nitrate | 37 (18%) |
| **Symptoms** |  |
| Typical AP | 79 (38%) |
| Atypical AP | 54 (26%) |
| Other | 78 (37%) |

Abbreviations: ACE, angiotensin converting enzyme; AP, angina pectoris; BMI, body mass index; CAD, coronary
artery disease; MI, myocardial infarction; PCI, percutaneous coronary intervention

**Table 2.** Flow, pressure and resistance indices stratified for FFR and PET findings.

|  | Overall | FFR+ PET+ | FFR+ PET- | FFR- PET- | FFR- PET+ | p value |
| --- | --- | --- | --- | --- | --- | --- |
| **[<sup>15</sup>O]H<sub>2</sub>O PET** |  |  |  |  |  |  |
| Regional hMBF<br>(ml/min/g) | 2.93 ± 1.17 | 1.76 ± 0.42 | 3.27 ± 0.49 | 3.75 ± 0.91 | 1.84 ± 0.32 | <0.01 |
| CFR | 2.91 ± 1.14 | 1.81 ± 0.58 | 3.24 ± 0.67 | 3.57 ± 1.03 | 2.09 ± 0.59 | <0.01 |
| MVR<br>(mmHg mL min/mL) | 26.80 ± 13.37 | 31.35 ± 13.96 | 19.50 ± 4.54 | 21.11 ± 5.12 | 43.62 ± 16.69 | <0.01 |
| **Invasive coronary angiography** |  |  |  |  |  |  |
| FFR | 0.89 (0.81 – 0.96) | 0.69 (0.55 – 0.75) | 0.77 (0.74 – 0.79) | 0.94 (0.88 – 0.99) | 0.91 (0.84 – 0.95) | <0.01 |
| QFR | 0.93 (0.80 – 0.99) | 0.70 (0.48 – 0.80) | 0.80 (0.70 – 0.89) | 0.98 (0.90 – 1.00) | 0.94 (0.87 – 0.98) | <0.01 |
| 3D QCA Diameter<br>stenosis (%) | 37.50 (23.00 – 49.00) | 57.15 (47.25 – 67.55) | 47.00 (39.75 – 55.65) | 33.00 (25.00 – 41.40) | 37.00 (32.50 – 47.00) | <0.01 |
| Angio-IMR | 27.74 (20.85 – 38.02) | 19.12 (15.10 – 29.50) | 24.18 (20.57 – 32.61) | 31.67 (23.47 – 40.63) | 28.19 (20.42 – 38.99) | <0.01 |

Abbreviations: FFR, fractional flow reserve; hMBF: hyperemic myocardial blood flow; IMR: index of microcirculatory resistance; MVR; microvascular resistance; PET, positron emission tomography; QCA, quantitative coronary angiography; QFR; quantitative flow ratio

**Figure 1.**
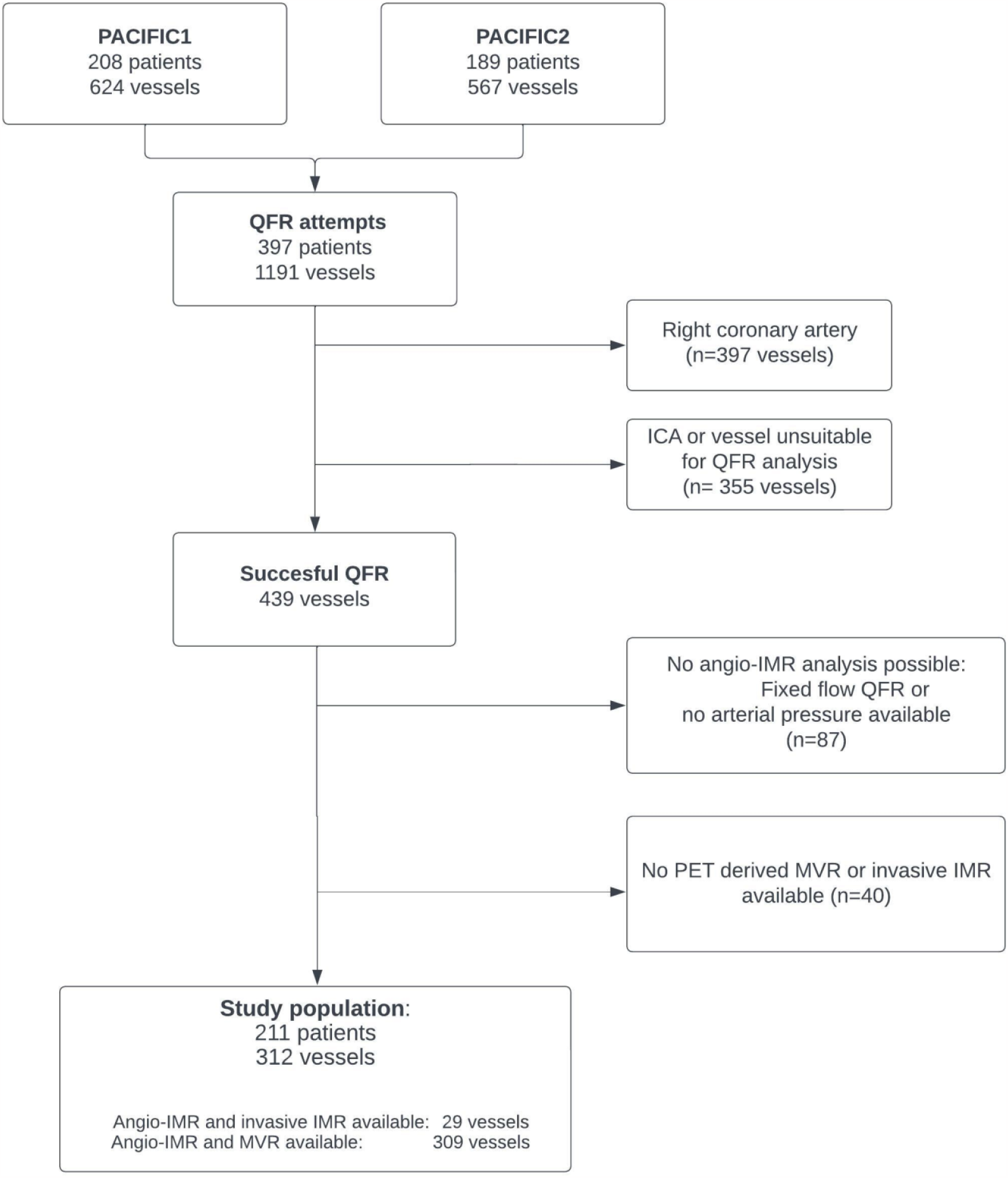
Study population outline.

### Validation of angio-IMR

A moderate correlation between angio-IMR and invasive IMR (r=0.48, p<0.01, figure 2) was observed. The Bland-Altman plot for agreement between angio-IMR and invasive IMR is shown in figure 2 (right). In patients with both angio-IMR and invasive IMR available higher median measurements of angio-IMR were found in comparison to invasively measured IMR (median 32.53, IQR 20.84 – 41.37 vs. 17.91, IQR 10.14 – 27.96). Angio-IMR was not related to MVR (r=-0.07, p=0.25; figure 3 left). Agreement between angio-IMR and MVR is summarized by a Bland-Altman analysis depicted in figure 3 right. After stratification for diameter stenosis the absence of correlation between angio-IMR and MVR was confirmed in vessels with ≥50% diameter stenosis (r=-0.13, p=0.32, n=66) or without (r=-0.02, p=0.77, n=243). Stratification for left anterior descending or ramus circumflex coronary arteries yielded similar results.

**Figure 2.**
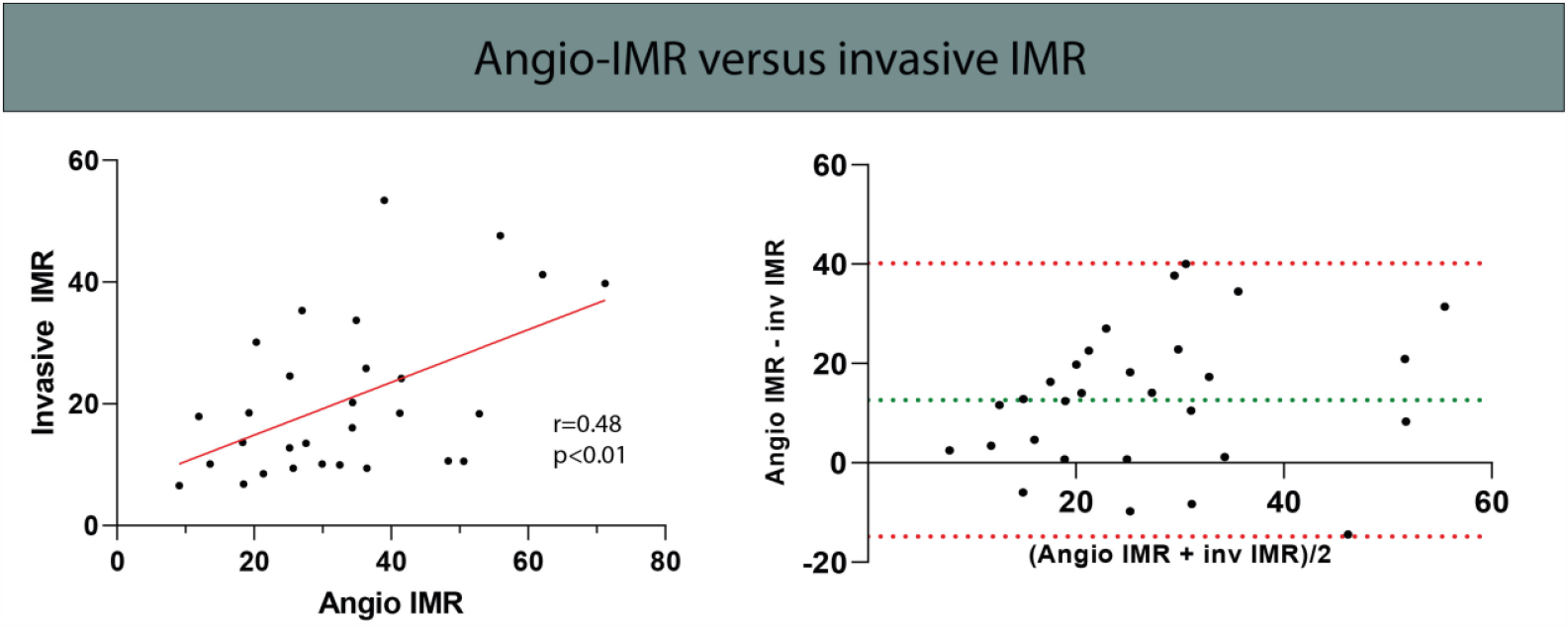
The correlation between angio-IMR and invasive resistance indices. Vessels with combined angio-IMR and invasively measured IMR were included only. Left: The correlation between invasive and angio-IMR. Right: Bland–Altman analysis summarizes concordance between angio-IMR and invasive IMR findings. Abbreviations: IMR, index of microcirculatory resistance

**Figure 3.**
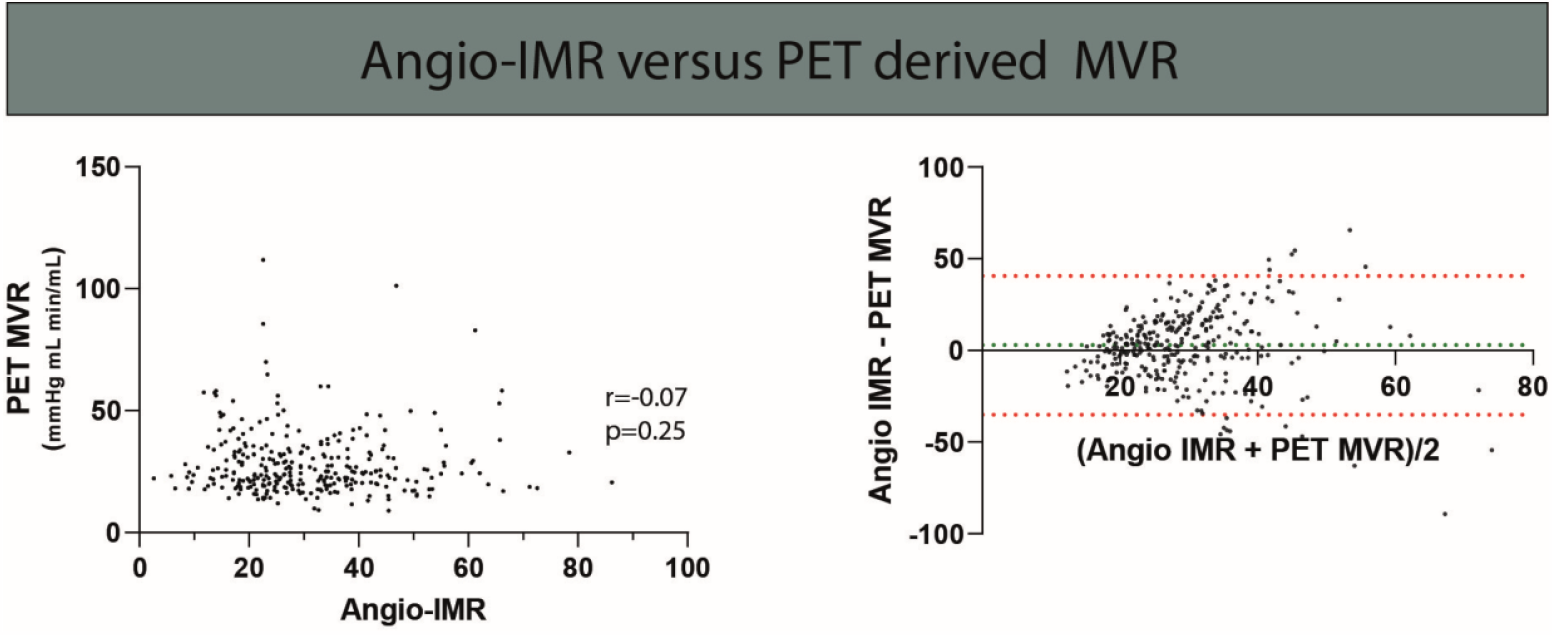
The correlation between angio-IMR and MVR. Vessels with combined angio-IMR and MVR were included only. Left: The correlation between angio-IMR and MVR. Right: Bland-Altman analysis summarizes concordance between angio-IMR and MVR findings. Abbreviations: IMR, index of microcirculatory resistance; MVR, microvascular resistance

### Angio-IMR in patients stratified for ischemia and obstructive CAD

Median vessel-specific angio-IMR values stratified for FFR and PET findings are shown in figure 4 (left). FFR negative vessels showed higher angio-IMR than FFR positive vessels. Vessels without obstructive CAD (FFR-) but with ischemia (PET+), the so called INOCA, showed higher angio-IMR levels than vessels with ischemia (PET+) and obstructive CAD (FFR+) (median 28.19 IQR 20.42 – 38.99 versus 19.12 IQR 15.10 – 29.50; p<0.01) but similar to vessels without obstructive CAD (FFR-) with a normal perfusion (PET-) (median 31.67 IQR 23.47 – 40.63; p=1.00). Similar findings were observed for vessels stratified among QFR and PET findings (figure 4 right). Vessels with a high angio-IMR but abnormal FFR are scarce (figure 4), and only 11 vessels were revascularized with angio-IMR in the highest quartile.

**Figure 4.**
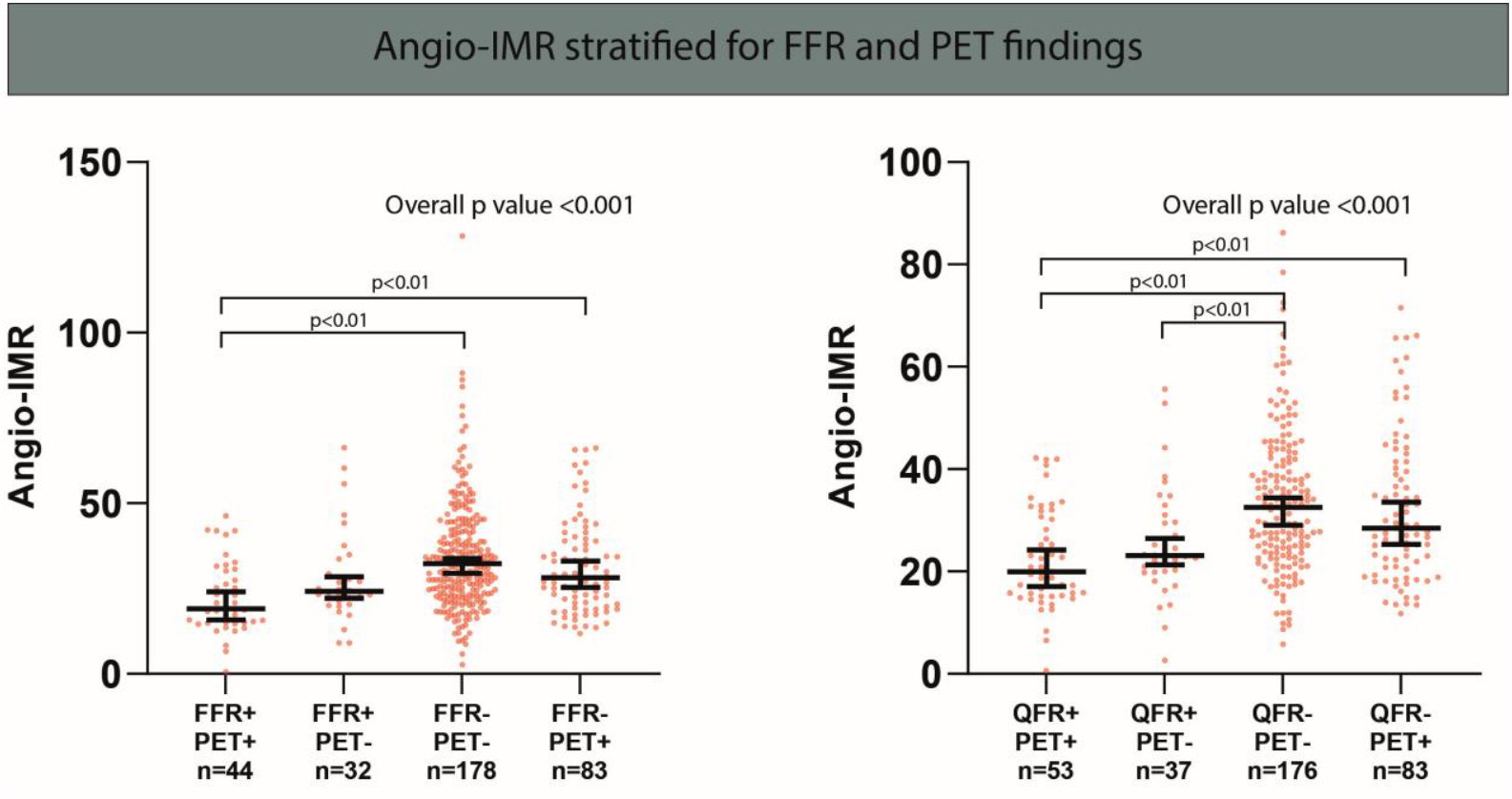
The Median (95% confidence interval) of angio-IMR stratified for PET and FFR (left) or QFR (right) results. Only significant p-values were displayed. Abbreviations: FFR, fractional flow reserve; IMR, index of microcirculatory resistance; PET, positron emission tomography.

## Discussion

In this investigator-initiated study we sought to validate angio-IMR and explore the potential of angio-IMR to detect INOCA in symptomatic patients. Our main findings are that we found a moderate correlation between angio-IMR and invasive IMR, whereas no correlation was observed between angio-IMR and PET derived MVR. Furthermore, angio-IMR was elevated in patients with non-obstructive CAD, irrespective for the presence of ischaemia, hampering the detection of INOCA. Our results question the validity of angio-IMR results and stress the importance of a state-of-the-art reference standard for the development of computer based algorithms to predict CMD.

An armamentarium of techniques to quantify invasive resistance are available, of which IMR and doppler-derived hyperemic microvascular resistance (HMR) are most well-known.(19) Both measures incorporate assumptions and it has been shown that invasive IMR and HMR cannot be considered equivalent.(20) Invasive IMR has practical advantages over HMR, e.g. simultaneous registration of epicardial and microcirculatory parameters, which lead to a more widespread use of invasive IMR.(19, 21) Our study confirms that angio-IMR and invasively measured IMR are, although moderately, significantly correlated. Of note, we found a slightly attenuated correlation between angio-IMR and invasively measured IMR compared to prior studies (r=0.70 and 0.76 vs. 0.48). (10, 22) The primary objective of our study was to validate angio-IMR against [^15^O]H_2_O derived MVR. Notwithstanding the moderate correlation between angio-IMR and invasively measured IMR we found no correlation between angio-IMR and MVR. Our findings question the value of the moderate relation between angio-IMR and invasively measured IMR and stress the importance of a state-of-the-art reference standard for the development of screening methods. Invasively measured IMR and PET derived MVR are not interchangeable indexes, e.g. invasive IMR is inversely associated with myocardial mass subtended by the coronary artery. (23) As CCTA was absent in the majority of patients we could not correct for subtended myocardial mass. To minimize the influence of myocardial mass we excluded the right coronary artery and placed the pressure and temperature sensors routinely on the same location in the distal third of the coronary. We chose PET derived MVR as reference standard because PET is a reliable method to quantify MBF. (24) Because hMBF during PET was not measured simultaneously with angio-IMR we corrected for driving pressure during PET by multiplying mean arterial pressure and FFR. The combination of these parameters measured at different points in time is valid since FFR is not dependent on hemodynamic variations, and hMBF and mean arterial pressure were measured simultaneously. Contrary, IMR as a reference standard has been criticized since it is difficult to obtain, operator dependent and, if collateral flow is not taken into account, non-specific to the microvasculature.(25-28) Hitherto there is no effective therapy for CMD and there are no reliable cut-offs to differentiate normal from abnormal MVR using PET. CAD and CMD share a complex interplay and a single reference standard might not capture this spectrum entirely. Presumably, invasive and non-invasive reference standards are complementary in understanding this complex interplay and the use of hard cut-offs will likely not improve our understanding of this pathophysiology.

The second objective of our study was to study whether angio-IMR could aid in identifying vessels with ischaemia but without obstructive CAD. In the cathlab, FFR assesses a coronary’s potential to improve in terms of perfusion after revascularization. It has been demonstrated by van de Hoef and colleagues that a high microvascular resistance masks FFR defined ischemia. (29) As such, we hypothesized that angio-IMR as a computed resistance index, could aid in the detection of these territories and guide appropriate treatment. We found a considerable group of INOCA vessels (FFR-, PET+) but since angio-IMR was elevated in vessels without obstructive CAD, irrespective of ischaemia status, angio-IMR did not aid in detecting INOCA.

Several PET flow tracers are available, but [^15^O]H_2_O most reliably quantifies myocardial flow because extraction of this freely diffusible tracer is complete, independent of flow and linearly related to myocardial perfusion.(30) Importantly, in a study by Kaufmann repeated measurements of MBF and CFR during the same study session were similar, demonstrating the validity of the technique.(24) Therefore, [^15^O]H_2_O PET is considered the golden standard for quantification of CFR and microvascular resistance. Our study results encourage the further development of angio-IMR. However, angio-IMR was not associated to the golden standard of resistance quantification, namely [^15^O]H_2_O PET derived MVR, and findings of angio-IMR should be interpreted with caution.

### Limitations

The results from the present study should be interpreted in consideration of some limitations. First, the majority of ICAs were not in adherence to a dedicated QFR protocol; therefore a significant number of vessels could not be included. Second, angio-IMR computation relies on contrast frame count. Since injection pressure was not standardized nor registered we could not correct for differences in injection pressures. Third, CCTA was not available in all patients. Therefore we could not correct for subtended myocardial mass. Fourth, although the [^15^O]H_2_O PET scans were analyzed for perfusion defects with a clinical judgement for vessel anatomy segmentation was based on standard coronary anatomy and individual anatomical variations cannot be excluded. It should be noted that individualized segmentation appears to have little impact on the diagnostic accuracy of PET. (31)

## Conclusion

Despite a moderate correlation between angio-IMR and invasively measured IMR, angio-IMR did not correlate with PET derived MVR and findings of angio-IMR should be interpreted with caution. Moreover, angio-IMR was elevated in vessels without obstructive CAD, irrespective of ischaemia status, hampering the identification of INOCA. The algorithm of angio-IMR should be further optimized against state-of-the-art reference standards.

## Data Availability

Data available on request from the authors

## Abbreviations

Angio-IMR: microcirculatory resistive index from functional angiography
CAD: coronary artery disease
CMD: coronary microvascular dysfunction
CMR: cardiac magnetic resonance
CFR: coronary flow reserve
FFR: fractional flow reserve
ICA: invasive coronary angiography
IMR: index of microcirculatory resistance
hMBF: hyperemic myocardial blood flow
HMR: hyperemic microvascular resistance
MVR: microvascular resistance
PCI: percutaneous coronary intervention
PET: positron emission tomography
QCA: quantitative coronary angiography

## Central illustration

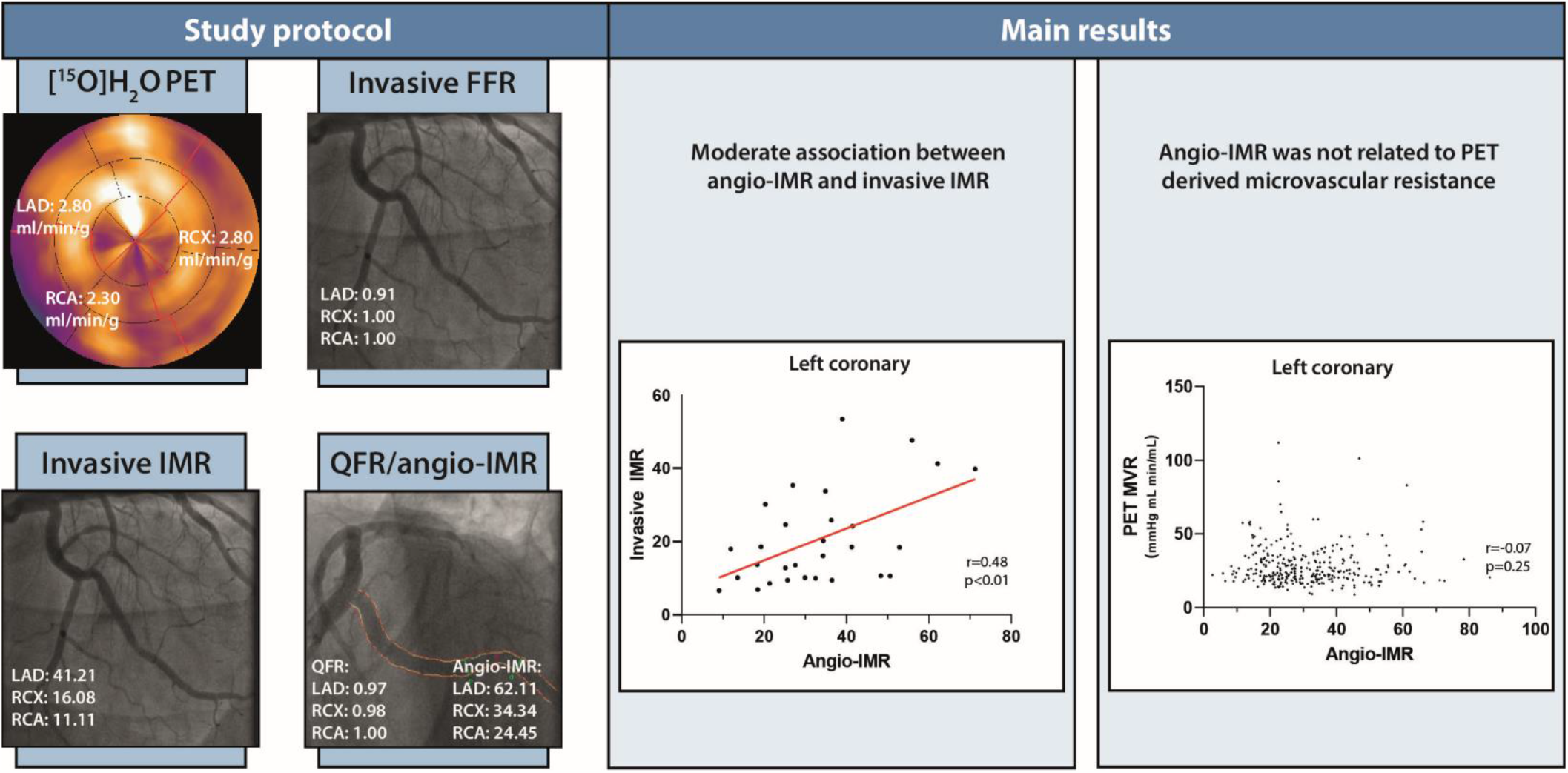

## References

1. Echavarria-Pinto M, Escaned J, Macias E, Medina M, Gonzalo N, Petraco R, et al. Disturbed coronary hemodynamics in vessels with intermediate stenoses evaluated with fractional flow reserve: a combined analysis of epicardial and microcirculatory involvement in ischemic heart disease. Circulation. 2013;128(24):2557–66.

2. Patel MR, Peterson ED, Dai D, Brennan JM, Redberg RF, Anderson HV, et al. Low diagnostic yield of elective coronary angiography. N Engl J Med. 2010;362(10):886–95.

3. Crea F, Bairey Merz CN, Beltrame JF, Berry C, Camici PG, Kaski JC, et al. Mechanisms and diagnostic evaluation of persistent or recurrent angina following percutaneous coronary revascularization. Eur Heart J. 2019;40(29):2455–62.

4. Knuuti J, Wijns W, Saraste A, Capodanno D, Barbato E, Funck-Brentano C, et al. 2019 ESC Guidelines for the diagnosis and management of chronic coronary syndromes. Eur Heart J. 2020;41(3):407–77.

5. Tonino PA, De Bruyne B, Pijls NH, Siebert U, Ikeno F, van’ tVeer M, et al. Fractional flow reserve versus angiography for guiding percutaneous coronary intervention. N Engl J Med. 2009;360(3):213–24.

6. Ford TJ, Stanley B, Good R, Rocchiccioli P, McEntegart M, Watkins S, et al. Stratified Medical Therapy Using Invasive Coronary Function Testing in Angina: The CorMicA Trial. Journal of the American College of Cardiology. 2018;72(23 Pt A):2841–55.

7. Song L, Xu B, Tu S, Guan C, Jin Z, Yu B, et al. 2-Year Outcomes of Angiographic Quantitative Flow Ratio-Guided Coronary Interventions. Journal of the American College of Cardiology. 2022;80(22):2089–101.

8. van Diemen PA, Driessen RS, Kooistra RA, Stuijfzand WJ, Raijmakers PG, Boellaard R, et al. Comparison Between the Performance of Quantitative Flow Ratio and Perfusion Imaging for Diagnosing Myocardial Ischemia. JACC Cardiovascular imaging. 2020;13(9):1976–85.

9. De Maria GL, Scarsini R, Shanmuganathan M, Kotronias RA, Terentes-Printzios D, Borlotti A, et al. Angiography-derived index of microcirculatory resistance as a novel, pressure-wire-free tool to assess coronary microcirculation in ST elevation myocardial infarction. The international journal of cardiovascular imaging. 2020;36(8):1395–406.

10. Mejia-Renteria H, Lee JM, Choi KH, Lee SH, Wang L, Kakuta T, et al. Coronary microcirculation assessment using functional angiography: Development of a wire-free method applicable to conventional coronary angiograms. Catheter Cardiovasc Interv. 2021;98(6):1027–37.

11. Danad I, Raijmakers PG, Driessen RS, Leipsic J, Raju R, Naoum C, et al. Comparison of Coronary CT Angiography, SPECT, PET, and Hybrid Imaging for Diagnosis of Ischemic Heart Disease Determined by Fractional Flow Reserve. JAMA Cardiol. 2017;2(10):1100–7.

12. Driessen RS, van Diemen PA, Raijmakers PG, Knuuti J, Maaniitty T, Underwood SR, et al. Functional stress imaging to predict abnormal coronary fractional flow reserve: the PACIFIC 2 study. Eur Heart J. 2022.

13. Danad I, Raijmakers PG, Harms HJ, Heymans MW, van Royen N, Lubberink M, et al. Impact of anatomical and functional severity of coronary atherosclerotic plaques on the transmural perfusion gradient: a [15O]H2O PET study. Eur Heart J. 2014;35(31):2094–105.

14. Danad I, Raijmakers PG, Driessen RS, Leipsic J, Raju R, Naoum C, et al. Comparison of Coronary CT Angiography, SPECT, PET, and Hybrid Imaging for Diagnosis of Ischemic Heart Disease Determined by Fractional Flow Reserve. JAMA Cardiol. 2017;2(10):1100–7.

15. Cerqueira MD, Weissman NJ, Dilsizian V, Jacobs AK, Kaul S, Laskey WK, et al. Standardized myocardial segmentation and nomenclature for tomographic imaging of the heart - A statement for healthcare professionals from the Cardiac Imaging Committee of the Council on Clinical Cardiology of the American Heart Association. Circulation. 2002;105(4):539–42.

16. Danad I, Uusitalo V, Kero T, Saraste A, Raijmakers PG, Lammertsma AA, et al. Quantitative assessment of myocardial perfusion in the detection of significant coronary artery disease: cutoff values and diagnostic accuracy of quantitative [(15)O]H2O PET imaging. Journal of the American College of Cardiology. 2014;64(14):1464–75.

17. Bom MJ, van Diemen PA, Driessen RS, Everaars H, Schumacher SP, Wijmenga JT, et al. Prognostic value of [15O]H2O positron emission tomography-derived global and regional myocardial perfusion. European heart journal cardiovascular Imaging. 2020;21(7):777–86.

18. van Diemen PA, de Winter RW, Schumacher SP, Everaars H, Bom MJ, Jukema RA, et al. The Diagnostic Performance of QFR and Perfusion Imaging in Patients with Prior Coronary Artery Disease. European heart journal cardiovascular Imaging. 2023.

19. Fearon WF, Kobayashi Y. Invasive Assessment of the Coronary Microvasculature: The Index of Microcirculatory Resistance. Circ Cardiovasc Interv. 2017;10(12).

20. Williams RP, de Waard GA, De Silva K, Lumley M, Asrress K, Arri S, et al. Doppler Versus Thermodilution-Derived Coronary Microvascular Resistance to Predict Coronary Microvascular Dysfunction in Patients With Acute Myocardial Infarction or Stable Angina Pectoris. Am J Cardiol. 2018;121(1):1–8.

21. Tebaldi M, Biscaglia S, Pecoraro A, Fineschi M, Campo G. Fractional flow reserve implementation in daily clinical practice: A European survey. International journal of cardiology. 2016;207:206–7.

22. Mejía-Rentería H, Wang L, Chipayo-Gonzales D, van de Hoef TP, Travieso A, Espejo C, et al. Angiography-derived assessment of coronary microcirculatory resistance in patients with suspected myocardial ischaemia and non-obstructive coronary arteries. EuroIntervention. 2022.

23. Echavarría-Pinto M, van de Hoef TP, Nijjer S, Gonzalo N, Nombela-Franco L, Ibañez B, et al. Influence of the amount of myocardium subtended to a coronary stenosis on the index of microcirculatory resistance. Implications for the invasive assessment of microcirculatory function in ischaemic heart disease. EuroIntervention. 2017;13(8):944–52.

24. Kaufmann PA, Gnecchi-Ruscone T, Yap JT, Rimoldi O, Camici PG. Assessment of the reproducibility of baseline and hyperemic myocardial blood flow measurements with 15O-labeled water and PET. J Nucl Med. 1999;40(11):1848–56.

25. Aarnoudse W, Fearon WF, Manoharan G, Geven M, van de Vosse F, Rutten M, et al. Epicardial stenosis severity does not affect minimal microcirculatory resistance. Circulation. 2004;110(15):2137–42.

26. Yong AS, Ho M, Shah MG, Ng MK, Fearon WF. Coronary microcirculatory resistance is independent of epicardial stenosis. Circ Cardiovasc Interv. 2012;5(1):103–8, s1-2.

27. Everaars H, de Waard GA, Driessen RS, Danad I, van de Ven PM, Raijmakers PG, et al. Doppler Flow Velocity and Thermodilution to Assess Coronary Flow Reserve: A Head-to-Head Comparison With [(15)O]H(2)O PET. JACC Cardiovascular interventions. 2018;11(20):2044–54.

28. Pijls NH, van Son JA, Kirkeeide RL, De Bruyne B, Gould KL. Experimental basis of determining maximum coronary, myocardial, and collateral blood flow by pressure measurements for assessing functional stenosis severity before and after percutaneous transluminal coronary angioplasty. Circulation. 1993;87(4):1354–67.

29. van de Hoef TP, Nolte F, EchavarrÍa-Pinto M, van Lavieren MA, Damman P, Chamuleau SA, et al. Impact of hyperaemic microvascular resistance on fractional flow reserve measurements in patients with stable coronary artery disease: insights from combined stenosis and microvascular resistance assessment. Heart (British Cardiac Society). 2014;100(12):951–9.

30. Driessen RS, Raijmakers PG, Stuijfzand WJ, Knaapen P. Myocardial perfusion imaging with PET. The international journal of cardiovascular imaging. 2017;33(7):1021–31.

31. Bom MJ, Schumacher SP, Driessen RS, Raijmakers PG, Everaars H, van Diemen PA, et al. Impact of individualized segmentation on diagnostic performance of quantitative positron emission tomography for haemodynamically significant coronary artery disease. European heart journal cardiovascular Imaging. 2019;20(5):525–32.

